# Strategic Purchasing Theory in Weak Governance Contexts: Evidence from Pakistan’s Sehat Sahulat Program

**DOI:** 10.64898/2026.03.21.26348956

**Authors:** Ahsanullah Khan Wazir, Amirhossein Takian, Shahzad Ali Khan, A. Akbari Sari, Seyed Mostafa Hosseini

## Abstract

Strategic purchasing theory, often centered in high-capacity institutions, is built on three main assumptions-purchaser autonomy, professional management, and stable governance. These assumptions are frequently unmet in low- and middle-income health systems, leading to questions about the theory’s applicability in settings with limited institutional capacity, contested authority, and devolved health governance. This study evaluates strategic purchasing and its theoretical bases within Pakistan’s Sehat Sahulat Program, a national health insurance initiative serving over 100 million beneficiaries.

A qualitative comparative case study utilized 22 interviews across five stakeholder groups, policy document reviews, and a four-rounded modified Delphi method with ten experts. Interviews were coded through hybrid deductive-inductive thematic analysis around Cashin and Gatome-Munyua’s six purchasing functions, achieving Cohen’s kappa 0.83.

The analysis revealed persistent deficits in six purchasing functions. benefit packages designed through political negotiation rather than health technology assessment; passive open empanelment with no provider deselection; flat-rate payments generating unchecked upcoding; performance monitoring capturing financial flows with zero clinical outcomes; a beneficiary registry last updated in 2011 excluding 30–40% of the urban poor; and nominal regulatory oversight without enforcement. Four institutional capacity constraints were identified including lack of statutory authority, human resource shortages, information system inadequacies, and coordination issues post-2010 devolution.

These findings represent precondition failure rather than implementation failure, a distinction with fundamental implications for how international evaluations attribute accountability and design remediation. A four-stage Strategic Purchasing Capacity Stages (SPCS) model is proposed, positioning Pakistan’s programme at Stage 1 (Administrative Purchasing) and demonstrating that prescriptions calibrated to Stage 4 capacity generate structurally determined failure regardless of financing. Strategic purchasing theory requires explicit specification of governance capacity thresholds below which its prescriptions become infeasible

**Key Messages:**

– Strategic purchasing is mis-aligned with most low- and middle-income countries’ health care delivery systems due to four institutional capacity weaknesses which are lack of purchasing authority, lack of specialized personnel, lack of data processing capacity, and failure to coordinate-an example being Pakistan’s Sehat Sahulat program.
– This study gives out specific details of a model called the Strategic Purchasing Capacity Stages (SPCS) Model that categorizes systems into Stage 1 through Stage 4 and identifies where Pakistan stands on this scale at Stage 1. Therefore, international agencies should always assess the system’s institutional capacity before recommending strategic purchasing reforms so that recommendations can be made to fit the system’s stage.

## Introduction

### The Strategic Purchasing Consensus and its Shortcomings

Countries in Low and Middle-Income Economies are urged to transition from passive to active or Strategic Purchasing as a means of achieving Universal Health Coverage (UHC) (1). Strategic purchasing was first developed by the United Kingdom’s National Health Service (NHS), to improve the use of public funds to achieve better population health outcomes, including benefit package design, provider selection, payment design, performance assessment, membership enrollment, and regulatory compliance (2) (3). The concept of strategic purchasing was developed in the context of high state capability whereas there is limited research that describes the necessary institutional arrangements (4). Seven countries within the RESYST Consortium were found to have gaps in their organizational capacity to engage in purchasing and, additionally, nine sub-Saharan African countries have some form of purchasing mandate through legislation yet lack even rudimentary purchasing capabilities (5) (6). What the theory currently lacks is an explicit specification of the governance capacity thresholds below which its six functions become structurally infeasible, a gap this study addresses through systematic analysis of Pakistan’s Sehat Sahulat Program. This research also examines how the post devolution governance system in Pakistan affects policy feasibility by using Kingdon’s Multiple Streams Framework (Kingdon 2011). This framework illustrates that the window for adopting new policies occurs when there is a convergence of problem recognition, available solution(s) and political receptiveness. In contrast to federal systems where authority is constitutionally contested, this paper asserts that this type of convergence must occur simultaneously at both the federal and provincial level, and therefore is a much more difficult requirement than single tier convergence and one that Pakistan’s institutional landscape has consistently thwarted.

#### Box 1

**Pakistan’s Institutional Landscape for Health Insurance: Contextual Parameters Relevant to Strategic Purchasing**

**Constitutional/Governance Structure**

The 18th amendment to Pakistan’s Constitution (2010) transferred the power of health planning and the delivery of services to four provinces and created a new federal coordination ministry that was greatly reorganized. The Federal Ministry of National Health Services can direct national-level health programs but cannot enforce compliance from provinces when they are dealing with purchasing failures caused by a lack of clear definition of federal versus provincial responsibility.

**Sehat Sahulat Program: Purpose**

The Sehat Sahulat Program (SSP), which began in 2015 and was expanded in 2019 and 2022, offers hospitalization benefits to all family members for an annual limit of PKR 1,000,000, providing coverage for over 100 million people in Pakistan who rely on public funds and the State Life Insurance Corporation. There are well documented deficiencies in the SSP, including:

– The use of the Benazir Income Support Program’s outdated poverty registry to determine eligibility does not include the urban poor
– The hospital-based design of the SSP leaves out outpatient and primary care
– There is no independent purchasing authority for the SSP, resulting in a lack of coordination and clarity among agencies working under the program
– Public spending for health in Pakistan is approximately 1% of its Gross Domestic Product, limiting further growth in health insurance.

**Study’s Contribution**

This research evaluates the SSP as a strategic purchasing mechanism utilizing the principles of strategic purchasing theory and the Cashin and Gatome-Munyua Framework to outline the requirements for implementing strategic purchasing effectively.

*Sources: Yang et al.* (*2024*)(*7*)*; Forman et al.* (*2022*)(*8*)*; Hasan et al.* (*2022*)(*9*)*; Pakistan Ministry of NHSR&C Status of Health Financing (2022)(10); WHO EMRO Pakistan Health Governance Profile* (*11*).

### Pakistan’s Sehat Sahulat Programme-A Boundary Condition Test for Strategic Purchasing Theory

Pakistan’s Sehat Sahulat Program is an ideal example for exploring limitations of strategic purchasing theory in healthcare. After the 18th Constitutional Amendment was passed in 2010, the federal government relinquished all of its health authority to the provinces without implementing a coordinated system, making it difficult to implement this program effectively (7, 12). Public expenditure on healthcare remains at approximately 1.0 percent of gross domestic product (GDP), among the lowest in the region and below the 5 percent recommended internationally for providing sufficient financial protection in healthcare (10). The Sehat Sahulat Program has been operational since 2015 and scaled up 2022. The objective of the program, to achieve universal health coverage via insurance covering up to PKR 1 million per family each year under a single insurer, remain greater than the capacity of the existing institutional infrastructure (8) (9).

Some of the major shortcomings of the Sehat Sahulat Program include the exclusion of outpatient services, the use of an inadequate poverty registry that fails to identify approximately 30-40 percent of households that are eligible for the program and there is no independent purchasing entity established for the program (11). Furthermore, the State Life Corporation’s role as both the provider and purchaser of insurance in the Sehat Sahulat Program creates a conflict of interest in terms of the corporation’s commercial mission and the program’s goals of improving public health. The governance structure of the Sehat Sahulat Program was created based on the 18th Amendment to the Constitution of Pakistan that emphasized the importance of provincial autonomy but failed to create the necessary purchasing capacity within the provinces.

### Study Aim and Theoretical Contribution

This research uses the Cashin and Gatome-Munyua framework for strategic purchasing to develop an enhanced Strategic Purchasing Capacity Stages model that identifies the gap between the institutional conditions of developing countries and the ability to engage in effective strategic purchasing. This research demonstrates that in order to create realistic policy models, international organizations should provide policy recommendations that take into account the current governance stage of the country and avoid recommending complex programs that may be beyond the capabilities of the country.

## Methods

### Study Design

The Sehat Sahulat Program (SSP) of Pakistan is examined using a qualitative comparative case study approach in order to identify the institutional requirements for successful strategic health purchasing. Because it identifies a phenomenon that has been heretofore unexplored, the program represents a revelatory case (4). The qualitative comparative case study design integrated three distinct data collection methods including primary qualitative interviews; programme documentation; and expert consensus. All three data streams were organized around the Cashin and Gatome-Munyua strategic purchasing progress tracking framework (4).

### Data Sources, Integration and Analysis

Twenty-two selected stakeholders participated in qualitative key informant interviews regarding the health insurance system in Pakistan. They represented government policy makers, program implementers, healthcare providers, and academic experts (see Table 2). Initially, federal level informants were interviewed first to provide context on SSP’s conceptual framework and subsequently, provincial representation from Sindh were interviewed to conduct federal-provincial analysis. The median number of years that the informants have worked in the health sector is 17 years and all informants are currently actively engaged in the design and implementation of SSP. Interviews lasted 45 to 75 minutes and were completed in either English or Urdu and were audio recorded with the participants’ consent. The Urdu interviews were transcribed, translated into English and de-identified so that no identifying information was present in the data used for the analysis.

The documents reviewed included, SSP Operational Framework, Provincial Health Benefit Packages for Islamabad and Sindh, Empanelment Criteria, Performance Reports 2019-2022, Parliamentary Discussions related to Financing for Health, and Provincial Development Plans related to Insurance Integration within the Health Sector. An interpretivist approach was selected for analysing these documents. Interpretivists are concerned with how people make sense of their experiences (Guba & Lincoln, 1985) (13).

To validate the findings from the study, a modified Delphi process involving ten expert panellists from federal and provincial levels of government was conducted. This process included four rounds with three rounds focused on analysis. In each round, panellists rated the institutional barriers to purchasing function deficits using Likert scale responses and provided open-ended rationale responses (7). Following each round, group ratings and comments were shared to facilitate further discussion. Consensus was established when there was at least 75% agreement or disagreement among the panellists regarding the specific item, and Kendall’s W > 0.7 was also used to establish strong consensus (14). The interview protocols were developed around the strategic purchasing framework functions (3) and an additional area was included to address federal/provincial coordination issues, which may be impacted by the 18th Amendment to the Constitution of Pakistan (see Table 1).

**Table 1.** – Interview Domain Framework: Purchasing Functions, Theoretical Sources, and Coding Approach.

| **Purchasing Function** | **Framework Source** | **Core Interview Questions** | **Coding Approach** |
| --- | --- | --- | --- |
| **1. Benefit Package Definition** | Cashin & Gatome-Munyua (2022) (4) | How are decisions made about which services the programme covers?<br>What evidence informs these decisions? Who has authority to change the package? | Deductive: evidence-based prioritization vs political determination;<br>inductive: emergent political economy themes |
| **2. Provider Selection & Network Development** | Cashin & Gatome-Munyua (2022) (4) | How are hospitals empaneled? What quality criteria apply? Can providers be de-listed for poor performance?<br>How are geographic gaps addressed? | Deductive: selective vs passive contracting;<br>quality assessment mechanisms;<br>inductive: power dynamics between PMU and providers |
| **3. Payment Mechanism Design** | Cashin (2015) (3); Mathauer et | How are payment rates determined and updated? Is risk adjustment applied? What evidence of moral | Deductive: rate-setting methodology, adjustment |
|  | al. (2017)<br>(6) | hazard or provider gaming exists? | mechanisms;<br>inductive: financial<br>sustainability<br>narratives |
| **4. Performance Monitoring** | Cashin & Gatome-Munyua (2022) (4) | What data are collected? How are data used to inform decisions? Is there outcomes monitoring or only utilization counts? Who receives performance reports? | Deductive: data-to-decision linkage; accountability mechanisms; inductive: information asymmetry themes |
| **5. Beneficiary Enrollment Management** | Govender et al. (2021) (5) | How are eligible households identified? What are the exclusion mechanisms? How current is the beneficiary database? How are urban poor populations reached? | Deductive: registry accuracy, exclusion patterns; inductive: socio-political determinants of enrollment targeting |
| **6. Regulatory Compliance Oversight** | Cashin & Gatome-Munyua (2022) (4) | Which body has statutory authority to oversee the programme? How are provider compliance and insurance company performance monitored? What enforcement mechanisms exist? | Deductive: statutory authority presence vs absence; enforcement capacity; inductive: accountability gap themes |
| **Federal-Provincial Coordination** | Nishtar et al. (2013) (12); Yang et al. (2024) (7) | How do federal and provincial authorities coordinate on programme decisions? What are the main friction points post-18th Amendment? Who has ultimate authority in disputes? | Deductive: H3 hypothesis testing (federal vs provincial positioning divergence); inductive: devolution constraint narratives |

*Protocol domains derived from Cashin and Gatome-Munyua (2022) with the addition of a federal-provincial coordination domain. Questions are abbreviated; full interview guide is available as Supplementary Material.*

An additional domain-federal-provincial coordination-was incorporated into the protocol framework to recognize that the 18th Amendment had created a governance structure that impacted all six purchasing functions. While the additional domain was not introduced during the analysis, it was pre-specified as an analytically essential component of the framework given the constitutional context of Pakistan and should be considered a theoretically motivated expansion of the framework rather than an example of post-hoc data fitting.

Data Integration used a Converging Triangulation Methodology to combine three types of data (Key Informant Interviews, Documentary Review, and Consensus Delphi). The Primary Themes found in Key Informant Interview data was corroborated through use of supporting documentation. The Delphi data supported all of the previous research findings with at least a 75% consensus among participants. All participant disagreements were noted specifically when looking at differences between the Federal and Provincial levels of government. The convergence of the Key Informant and Delphi data indicated that there was strong evidence to support previous findings. The divergence of these two sets of data suggested that there were very different Institutional Realities at each level of government. The Documentary Analysis was completed to provide evidence to support previous claims and to provide additional context to help explain why things happened as they did. A high degree of Explanatory Completeness was achieved by using this method, but no Formal Weighting was applied across methodologies.

### Analytical procedures

An analytical process of deductive coding (Braun & Clarke, 2006) (15) using an established framework of 6 stages, was used to apply a pre-established set of deductive codes drawn from 7 protocol areas (addressing six purchasing functions and federal/provincial coordination), which were used consistently across 22 transcripts to identify deficits. Simultaneously, an inductive coding process was employed to reveal unforeseen themes associated with the political economy of program development and institutional constraints, ultimately yielding four capacity constraints common to both informant groups and governance levels. A systematic process of intercoder reliability was achieved through two independent analysts conducting the coding of a randomly selected 20% sample, achieving a Cohen’s Kappa of 0.83, thus indicating a high degree of agreement. The coding process was systematized and discrepancies were addressed collaboratively, creating a single unified codebook for the dataset.

### Ethical considerations and rigor

Ethics approval was granted for this study from the Institutional Review Board of Tehran University of Medical Sciences (IR.TUMS.SPH.REC.1400.187) and the Ethics Review Committee of Health Services Academy Pakistan (HSA/ERC/23/008). It is consistent with the Declaration of Helsinki and the principles outlined in the Belmont Report (16). All participants provided informed consent for their participation in the study, thereby ensuring they participated voluntarily and had the opportunity to maintain confidentiality as well as withdraw from the study at no cost or penalty. Strategies were adopted for establishing trustworthiness including credibility through the researcher’s extensive engagement with the research site, member checking with senior informants, and the use of triangulation methods to validate the findings. Transferability was also enhanced by providing a detailed description of the research context.

To minimize potential confirmation bias, the researcher has expertise in health policy in Pakistan and utilized a deductive coding approach to ensure neutrality. Additionally, the study did not receive funding from outside sources, therefore, the results are free from the influence of stakeholders and add to the overall validity of the analysis (Table 2).

**Table 2.**
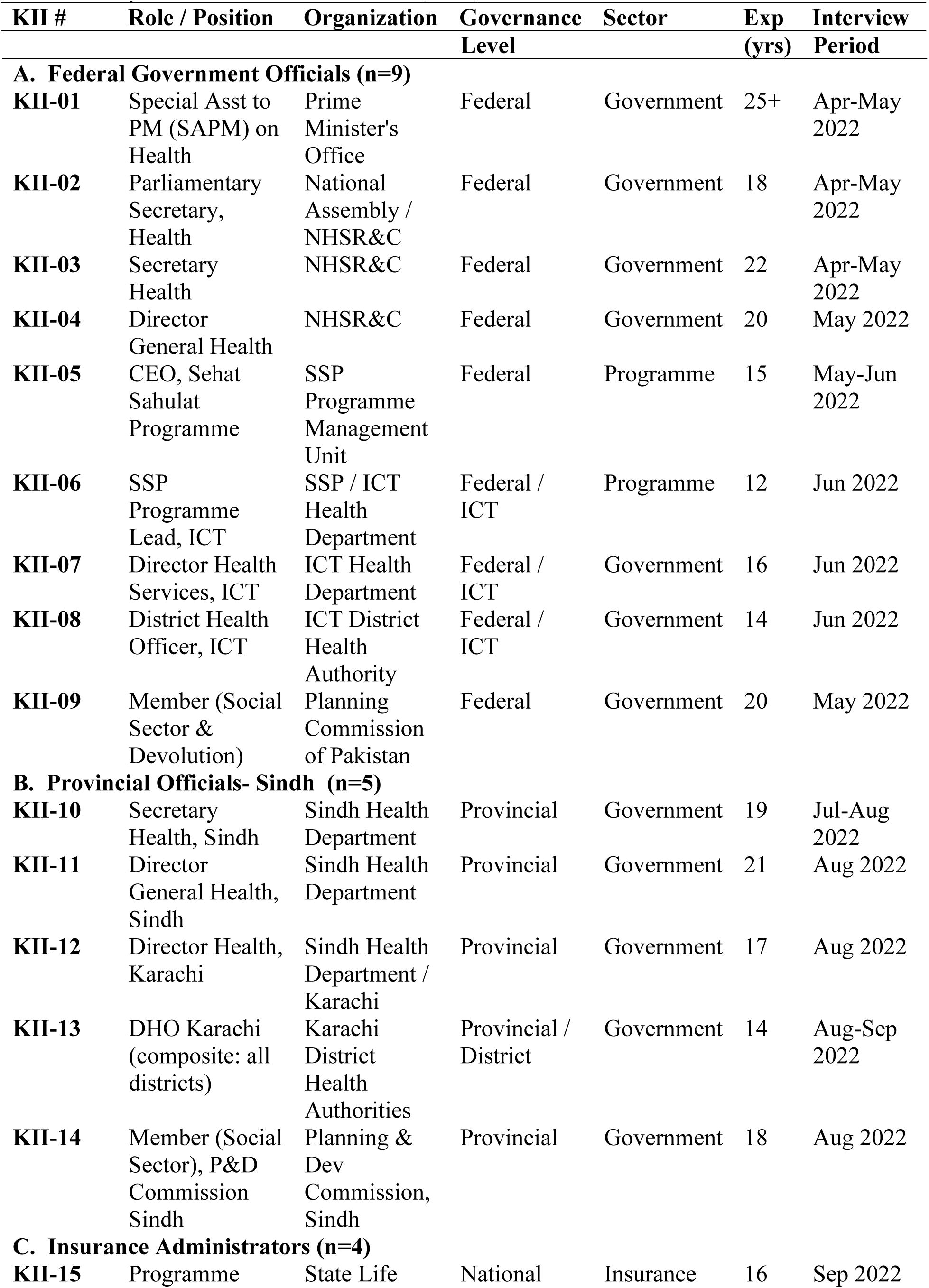
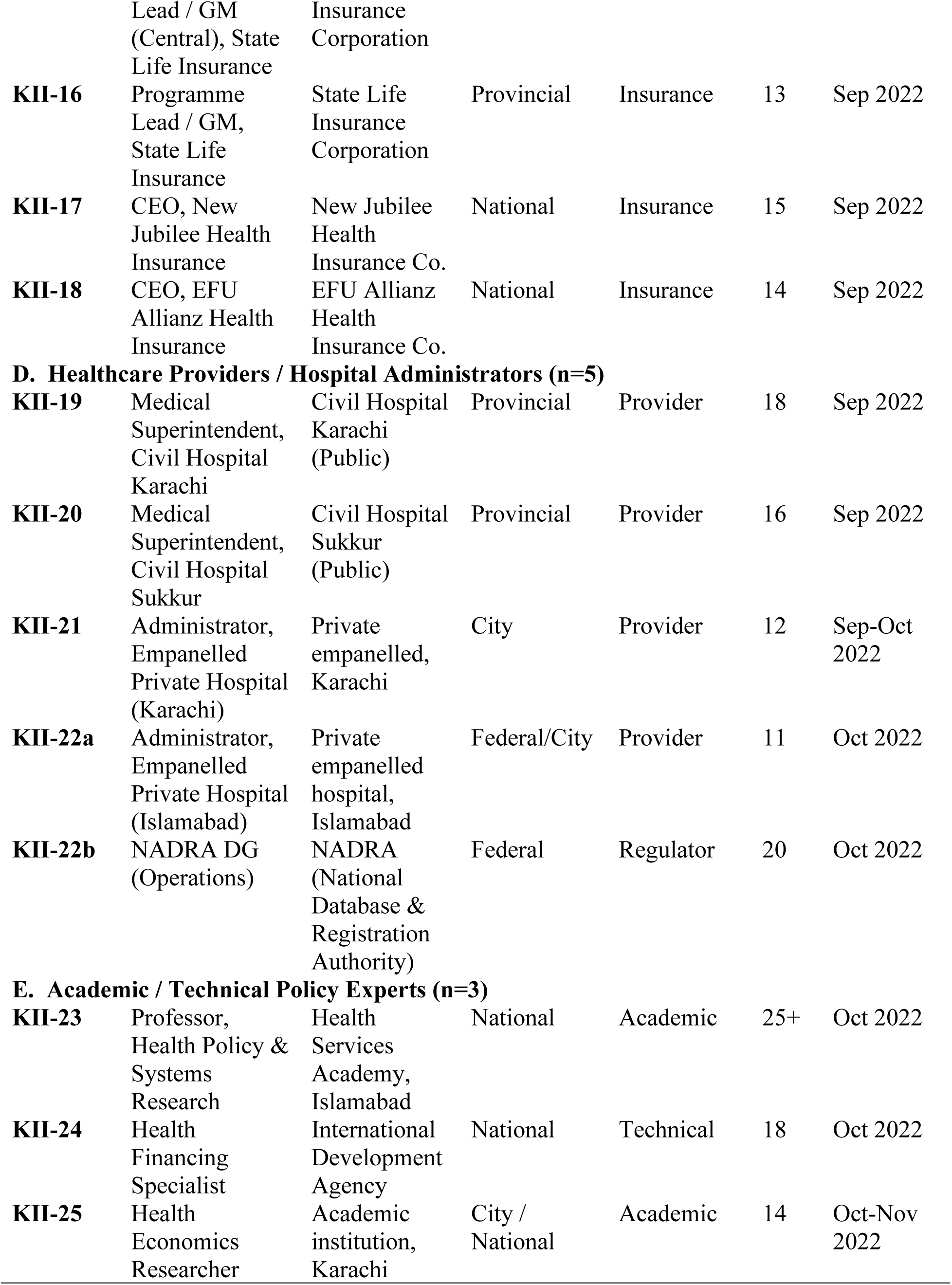
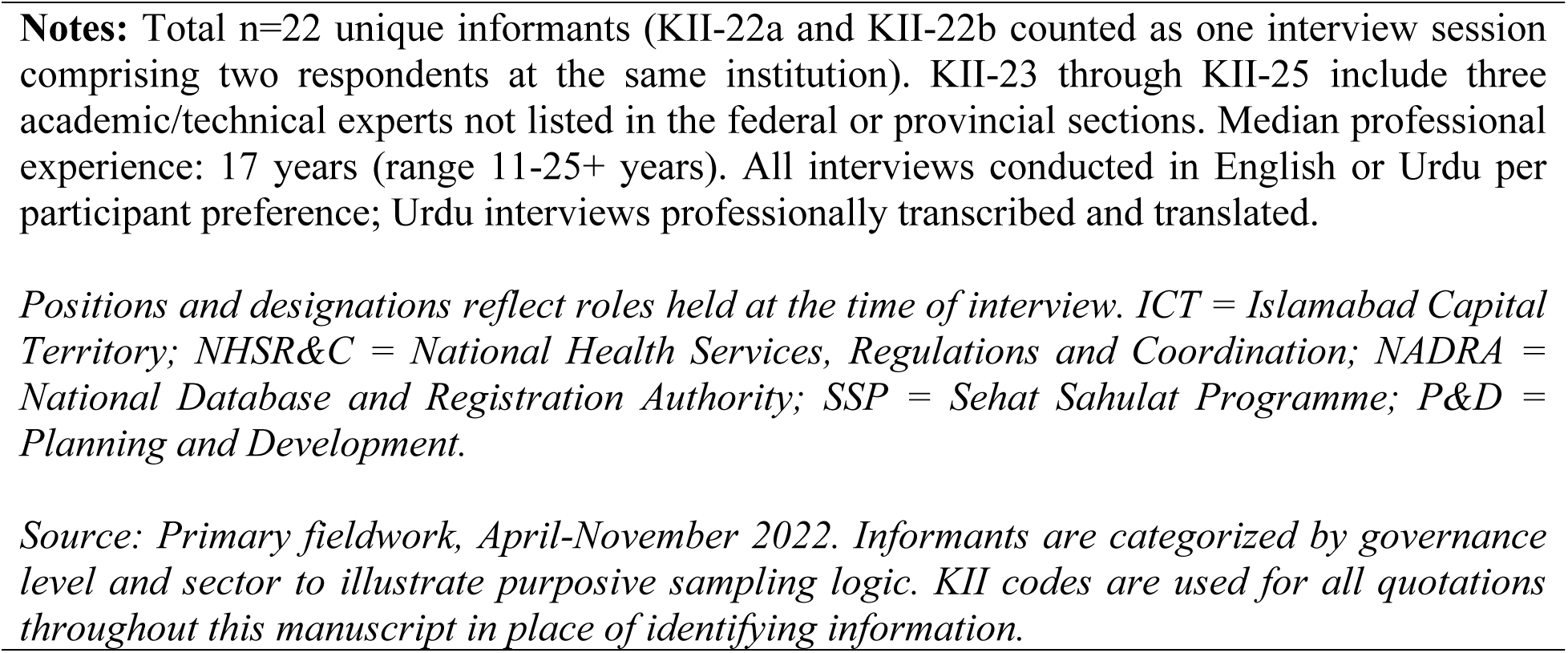
Key Informant Characteristics (n=22)

## Results

The thematic analysis revealed that there are consistent failures in each of the six strategic purchasing functions outlined by Cashin and Gatome-Munyua’s framework. The four functions with the greatest shortfall also had the most dysfunction, the two remaining functions were virtually non-functional. The four institutional capacity constraints were found to be structural barriers hindering effective purchasing, as described in detail in Tables 3 and 4.

**Table 3.**
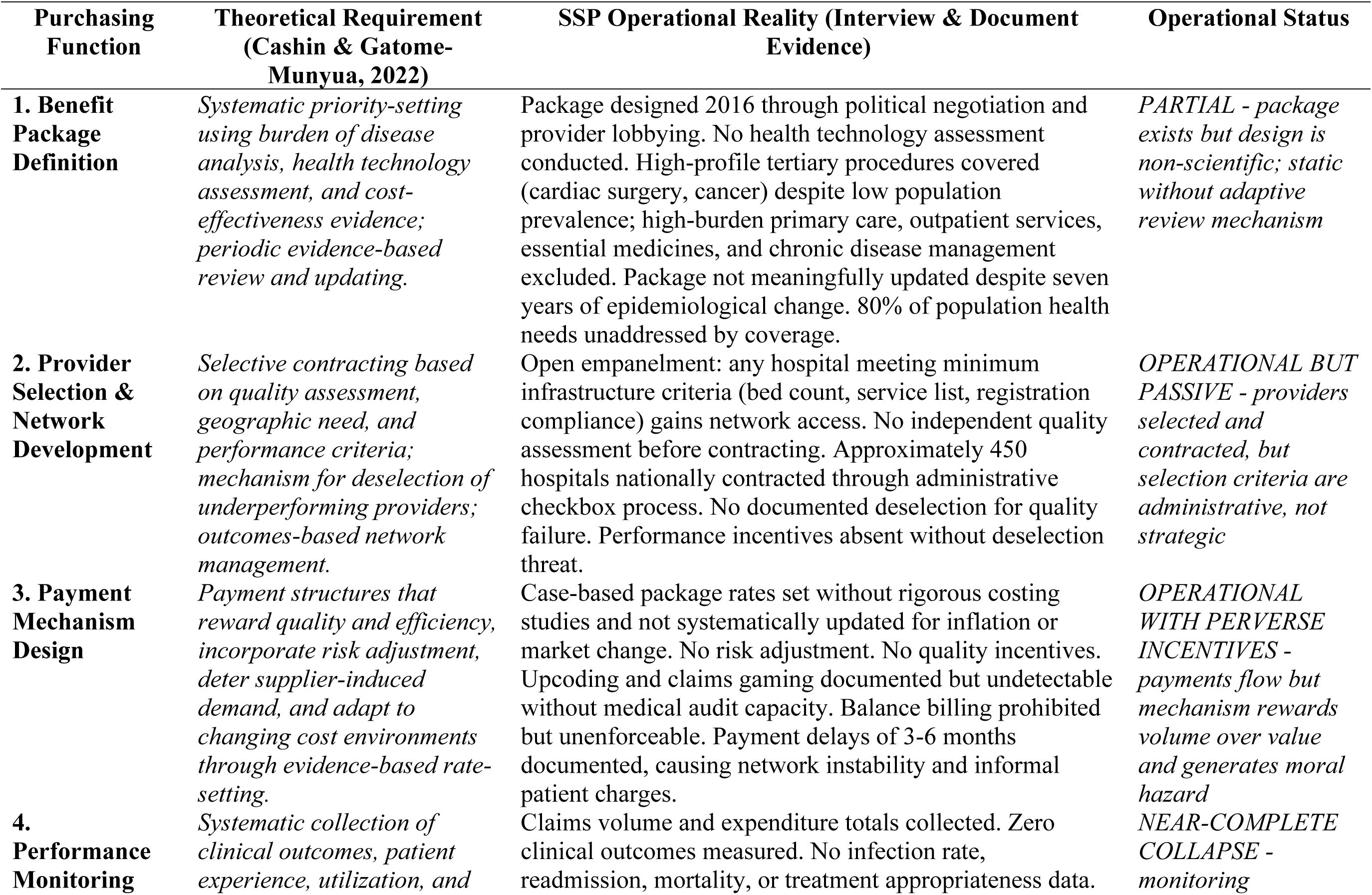
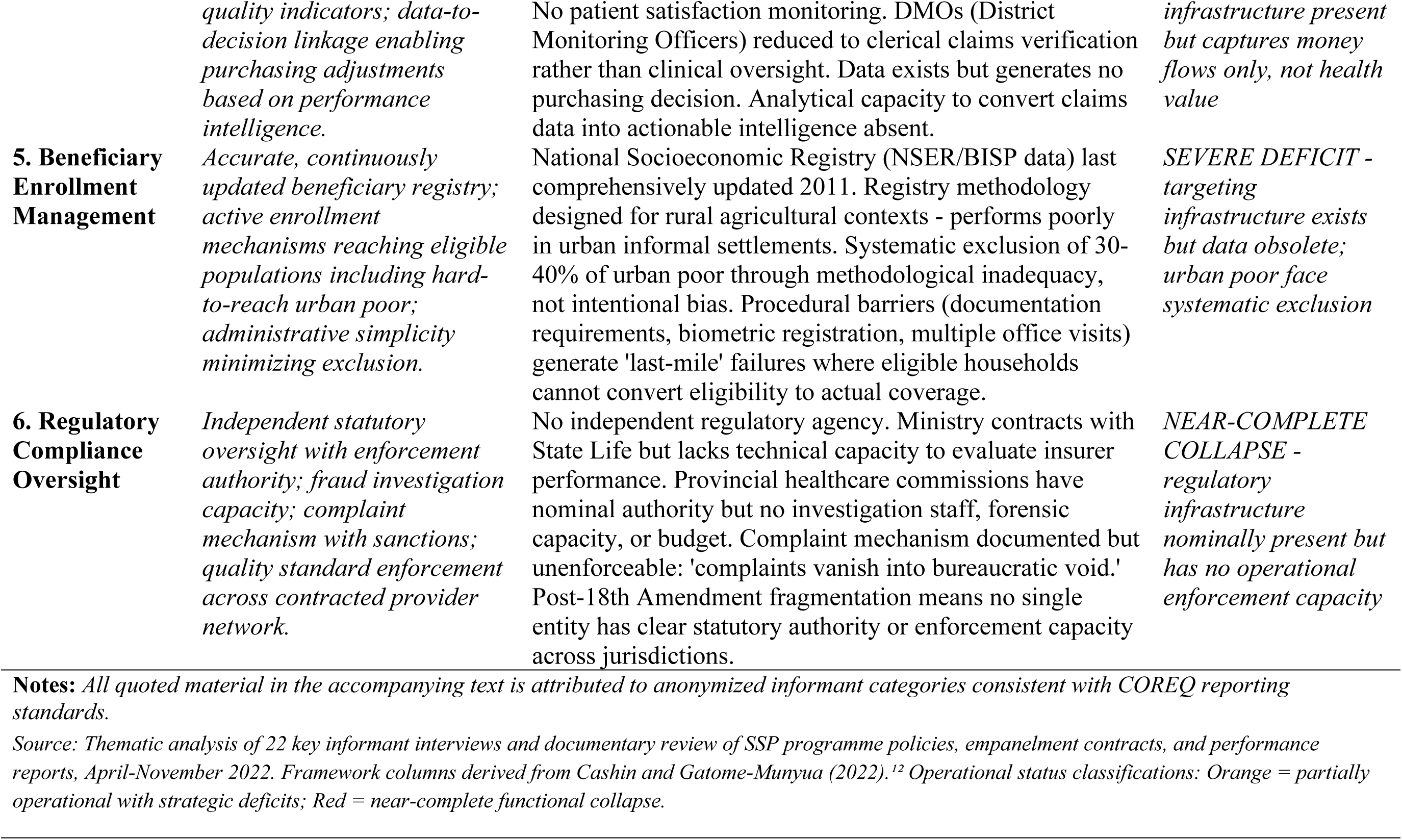
Strategic Purchasing Functions: Theoretical Requirements versus SSP Operational Reality.

**Table 4.**
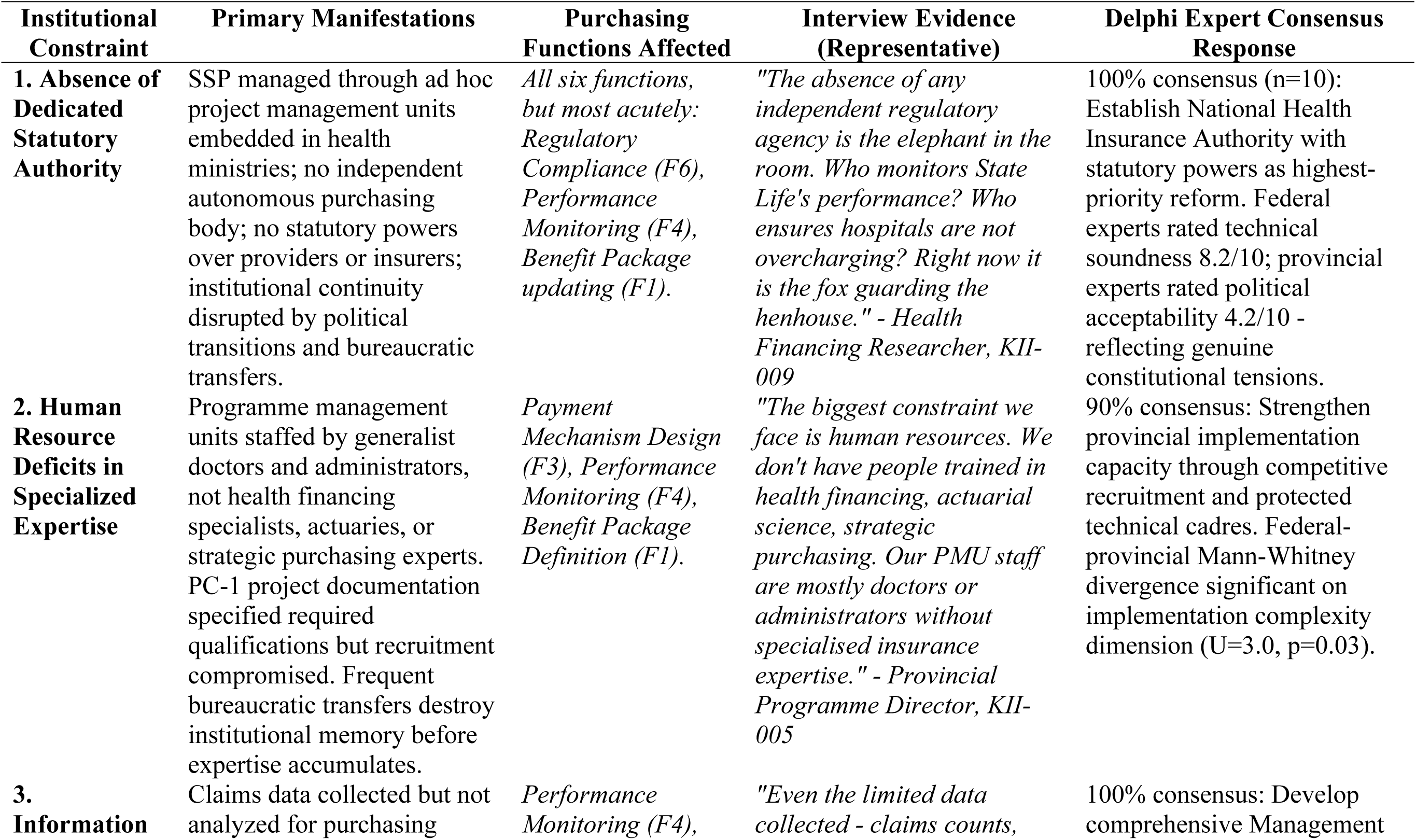
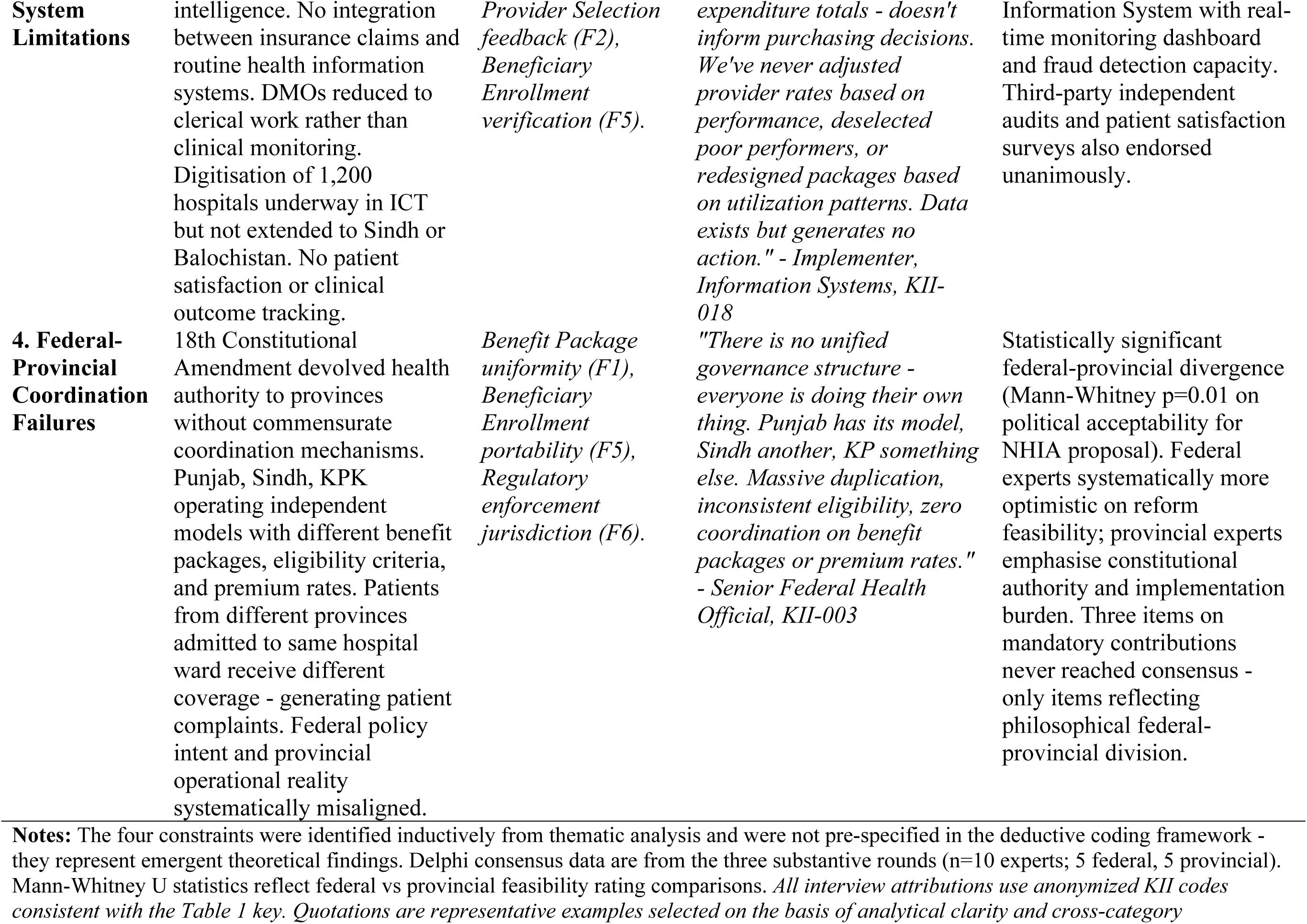
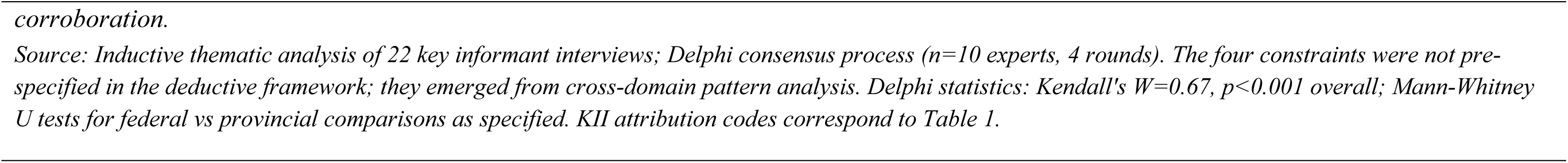
Four Institutional Capacity Constraints: Evidence Matrix.

### Benefit Package Determination: Politics Supplanting Evidence

The SSP benefit package has a wide variety of (over 1,000) medical treatments that can be used to treat many different health issues. However, the processes used to add these treatments have been piecemeal i.e., they did not evolve as part of an overarching plan. Procedures with political visibility are more likely to be added, whereas there is little likelihood of adding treatments for chronic diseases because of the complexity of managing them. The decision-making process for what would be included had no logical basis related to their cost, prevalence, or effectiveness. Simply put, it was a matter of negotiation.

*“The benefit package developed through negotiations with providers and politics rather than through health technology assessments or burden of disease analyses. Cardiac surgery, although it has a relatively low prevalence among the population, received coverage due to its visibility whereas diabetes management, despite being a high-burden condition, remains outside of the benefit package since it is chronic rather than acute.”*

*– Federal Government Official, Strategic Purchasing Unit, KII-01*

The failure of health insurance plans to provide adequate coverage for low-income urban residents’ health care needs limits access to many health-related services. In particular, approximately 80 percent of all health care provided to this population which includes primary care visits, outpatient care and chronic disease management that are not covered by the plan’s benefits package. As a result, the bulk of the plan focuses on providing coverage for infrequent hospitalizations (which may be associated with high medical bills), while leaving individuals to pay out-of-pocket for more frequent episodes of illness. Furthermore, since the benefits package was adopted in 2016, it does not account for changes in either disease prevalence or the cost of delivering health care; therefore, it has become outdated.

### Selection of Providers: Open Empanelment as a Form of Passive Purchasing

Once a hospital is on the network it cannot be removed from it (regardless of quality of care, etc.) which provides little incentive for performance as contract templates focus on infrastructure versus what can be measured, therefore further proving that the process is an entirely passive one. The current contracts are simply empanelment agreements with no provisions for escalating noncompliance issues. In essence, the contracting process is simply an administrative process, as one government official who is responsible for contracting for the program stated, “we don’t assess the quality of any hospital before we contract with them. We track no outcomes after we contract with them. We compare no hospital’s performance with other hospitals’ performance. Our process is purely administrative.”

*“Any hospital meeting minimal infrastructure criteria-certain bed count, some specialized services, registration compliance-qualifies for empanelment. We conduct no independent quality assessment, outcome tracking, or performance evaluation before contracting. Selection is an administrative checkbox exercise rather than a strategic quality-based choice.”*

*– Government Official, Contracting Unit, KII-05*

### Payment Mechanisms: Flat Rates Without Intelligence

A “flawed” Case-Based Payment System (CBPS) is used by the SSP in which all hospitals receive the same amount of reimbursement money for each treatment provided and this reimbursement money does not account for inflationary pressures, nor do they have the ability to provide differential payments based on varying levels of quality among different providers. As a result, hospitals will continue to cut costs as much as possible while providing lower quality care, and hospitals will continue to engage in the practice of “upcoding,” and there is currently no mechanism available to audit claims to stop such abuse.

*“Service providers alter coding to obtain the largest possible reimbursement amount for each claim submitted. Service providers will bill for a higher level of care than actually provided. Since the SSP Program has no utilization review or medical audit capabilities, we cannot detect patterns of abuse and manipulation by service providers. Therefore, service providers understand that they can manipulate their billing practices to optimize revenue instead of optimizing quality of care.”*

*–Government Official, Finance Department, KII-02*

Delayed payment for claims is a major operational weakness, with providers experiencing delays of 3 to 6 months before they receive payments from reimbursements. In addition to this, federal regulations prohibit providers from asking patients for co-payments; however, providers are unable to reliably anticipate when they will be paid by the program and therefore sometimes require patients to make upfront payments. These forms of payment contribute to undermining the financial protection commitment made by the program on behalf of its patient population.

### Performance Monitoring: Data Collection vs. Utilization

The SSP Program has developed extensive data collection systems for claims, expenditures, and procedures; however, it has not been successful in converting that data into useful purchasing information (purchasing intelligence) for decision-making.

*“We track claims volume and spend, but we measure zero clinical outcomes. We cannot tell if purchased services produce better health outcomes, reduce complications, or meet the needs of our customers. Our monitoring system measures money flows, not value creation.”*

*–Government Official, Monitoring and Evaluation, KII-04*

The district health department does not collect any clinical quality indicators, such as infection or hospital readmission rates, nor does it gather patient satisfaction data. The role of monitoring officers at the district level has diminished significantly; instead of being involved in the clinical management of care, they have become largely responsible for clerical/administrative work. Thus, the primary function of the monitoring system is that of a financial audit, with no incentive for providers to improve their performance (i.e., changes to reimbursement) and no providers have been removed from the program nor has anyone’s benefit package changed as a result of their performance on the program.

### Beneficiary Enrollment: Systematically Excluding Eligible Populations Through Methodology

Eligibility determination for the SSP Program is based on the National Socioeconomic Registry-the same database that the Benazir Income Support Program uses to identify households that qualify for cash transfers due to their poverty status. The National Socioeconomic Registry was last comprehensively updated in 2011. In Karachi, where the urban population has grown substantially and household circumstances have changed considerably during the past decade, the eligibility database for the SSP Program represents a historical record of income and wealth status, not a current record.

*“The National Socioeconomic Registry used to determine eligibility has not been comprehensively updated since 2011. Rapidly urbanizing areas like Karachi render targeting data obsolete, thereby systematically excluding new poor while including households that experienced improvements in their economic circumstances.”*

*– Government Official, Eligibility Determination, KII-03*

The approach taken by the SSP program to determine which populations will be targeted is inadequate, based upon out-of-date rural criteria that do not function well in urban informal communities. This results in 30-40% exclusion of the urban poor. Eligible household members often experience difficulties with documentation, biometric registration, and lengthy time frames associated with the application process. As such, many are denied insurance coverage, although technically eligible. Thus, the program may have the appearance of being universal but is in fact exclusionary.

### Regulatory Compliance Oversight: Symbolic vs. Substantive

There is also insufficient independent regulatory supervision. The Ministry of National Health Services does not possess the necessary technical capabilities to review the administration of the program by the State Life Insurance Corporation. Provincial health care commissions also do not have sufficient resources or capability to provide meaningful oversight to their contracted hospital providers.

*“There is no independent regulatory body overseeing the SSP Program. Who oversees State Life’s performance? Who prevents hospitals from over-charging? Who protects the rights of patients when claims are denied? At present, it is the fox guarding the hen house -- the Ministry contracts with State Life to operate the SSP Program but has no technical capability to evaluate whether State Life is operating the program effectively or ineffectively.”*

*– Health Financing Researcher, KII-009*

The ability to complain about a patient’s experience or problem is an option under this system; however, when it comes to the Ministry enforcing the resolution of those complaints, there appears to be a significant issue. When you add the fact that there is fragmentation among numerous agencies with ambiguous roles and responsibilities, the possibility of collaboration and accountability is severely reduced. Therefore, the regulatory oversight of the SSP Program has been unsuccessful.

### Four Institutional Capacity Constraints Underlying Functional Deficits

Inductive analysis using 22 Key Informant Interviews found four structural barriers, typical to all stakeholders, and at both the federal and provincial level, limiting the ability to implement purchasing functions regardless of program intent or financial support.

#### Lack of an Independent Statutory Purchasing Authority

The SSP uses a temporary Project Management Unit (PMU), operating out of the Ministry of Health, and does not possess an independent statutory basis, decision making authority, or the power to legally sanction providers or require them to disclose data. A Health Financing Researcher noted:

*“ There is no independent regulatory body overseeing the SSP Program. Who will oversee State Life’s performance? Who will stop hospitals from over-charging? It is currently the fox guarding the hen house.”*

Delphi Consensus (n = 10) ranked establishment of an independent statutory-based National Health Insurance Authority (NHIA) as the top priority for reform, although federal respondents gave political feasibility a rating of 8.2/10 and provincial respondents a rating of 4.2/10. This highlights the existing constitutional tension outlined in section 4.3.

#### Human Resources Shortage in Specialized Expertise

Strategic purchasing involves having personnel with specialized expertise in Actuarial Science for Benefit Costing, Health Economics for Incentive Design, Informatics Skills for Performance Analytics, and Contract Management Competency - none of which exist within SSP program units.

*“Our biggest problem is our human resources. We do not have the required training in Health Finance, Actuarial Science, or Strategic Purchasing. All of our PMU staff are primarily medical professionals or Administrators who lack specialized insurance knowledge” (Provincial Programme Director, KII-005).*

This shortage is exacerbated by the constant transfer of government employees resulting in a lack of institutional memory.

Delphi Consensus (n = 10) agreed (90%) that the best way to strengthen the capacity for implementing the Provincial Programs would be through the competitive hiring process, although there were significant differences between federal and provincial respondents regarding how complex the programs were to implement (Mann-Whitney U = 3.0, p = .03).

#### Limitations in Information Systems

The Information Infrastructure of SSP can collect data on claims volumes and expenditures, however, the analytical architecture necessary to transform transactional data into purchasing intelligence is absent.

*“The limited data that we collect - claims volume, total expenditure, etc. - is never used to make purchasing decisions. To date, we have not modified provider payment rates based on performance, removed poor performing providers, nor re-designed packages based on utilization trends. The data exists, but takes no action” (Implementer, Information Systems, KII-018).*

Digitalization of 1200 hospitals in the Islamabad Capital Territory is underway, however, digitalization of Sindh or Balochistan has not been implemented.

Delphi Consensus (n = 10) was unanimous (100%) that a comprehensive management information system with the capability to provide real-time monitoring and detect fraudulent activity would be needed to support the purchasing function.

#### Failures in Federal-Provincial Coordination

The 2010 devolution of authority related to health to the provinces did not include mechanisms to coordinate these new authorities, creating an environment of separate, non-compatible programs.

*“There is no single governing structure - everybody is doing their own thing. Punjab has its model, Sindh has another, KP has something else. There is massive duplication of effort, inconsistent eligibility criteria, and no coordination on what benefits to offer, nor on premium rates” (Senior Federal Health Official, KII-003).*

Families moving from province to province lose coverage portability; Patients located in adjacent hospital beds may receive completely different services under completely different provincial schemes. The statistically significant difference between federal and provincial respondents in terms of the perceived political acceptability of establishing an independent statutory-based NHIA (Mann-Whitney U = 2.0, p = 0.01) supports the view that the coordination failure is reflective of differing institutional positions rather than communication failures that could be resolved administratively.

## Discussion

### Strategic Purchasing Theory and its Embedded Institutional Assumptions

The development of strategic purchasing theory was based on the analysis of systems like Thailand, Estonia, and New Zealand. These systems had an existing administrative infrastructure, a professional technical capacity, and an institutional arrangement that was stable and operational prior to the articulation of their purchasing ambitions (4, 17). Cashin and Gatome-Munyua identify the functional requirements for strategic purchasing, however, they did not establish the governance capacity thresholds below which these functions would be structurally infeasible. Consequently, this omission has resulted in a substantial misdiagnosis in which failures resulting from the absence of requisite institutional conditions are incorrectly attributed to inadequate implementation, leading to remedies addressing the incorrect issue. The results of this research provide a basis for making this distinction concrete that what Pakistan’s SSP demonstrates is not a failure to implement strategic purchasing, but a failure of the structural conditions necessary for implementing strategic purchasing

The results show that strategic purchasing can be viewed in terms of Institutional Maturity, which leads to the development of the Strategic Purchasing Capacity Stages (SPCS) Model (Table 5). As given below, this model outlines how sophisticated purchasing practices are measured against governance capacity thresholds and recommends that policy recommendations be aligned with the current capacity of a country to purchase strategically, as opposed to an ideal capacity.

– Administrative Purchasing, Stage 1, is characterized by Systems that primarily function as financial intermediaries with benefits negotiated politically as opposed to being evidence-based. Actuarial foundations for payment systems do not exist and therefore lead to perverse incentives without the capability to monitor these activities. An example of a country at this level is Pakistan’s Sehat Sahulat Program, which is representative of the ceiling for Low- and Middle-Income Countries (18).
– Selective Purchasing, Stage 2, occurs when purchaser decision-making is informed by the quality of services provided by providers. Quality criteria evolve to include service quality indicators and enable purchasers to remove non-compliant providers from consideration for contract awards. This level of activity requires both foundational health information systems and regulatory authority to ensure compliance.
– Performance Purchasing, Stage 3, begins when purchaser decision-making is influenced by the outcomes achieved by providers. Providers who achieve high quality receive larger contracts; accordingly, there must be a purchaser body with the capacity to independently analyze data and provide coordination across multiple levels of government.
– Full Strategic Purchasing, Stage 4, represents the ideal where all functions operate effectively. This is best exemplified by Thailand’s National Health Security Office. At this level, evidence-based benefit design, quality incentives and strong regulatory frameworks are present(4, 19).

**Table 5.** Strategic Purchasing Capacity Stages (SPCS) Model.

| Stage | Operational Component/s | Missing Component/s | Country Examples | Key Requirement for Progression |
| --- | --- | --- | --- | --- |
| Stage 1<br>Administrative | Basic claims registration; benefit package set politically; all willing providers enrolled; flat-rate payment | No quality criteria; no outcomes data; no performance accountability; no regulatory capacity | Pakistan SSP (current); many sub-Saharan fragile states; early-stage LMIC programmes | Establish dedicated statutory purchasing authority; begin building health information infrastructure |
| Stage 2<br>Selective | Quality-based provider selection begins; basic utilisation data collected; empanelment criteria tightened | Performance does not yet feed payment; regulatory authority limited; beneficiary databases incomplete | Ghana NHIS early phase; some East African community health insurance schemes | Develop evidence-based empanelment criteria; invest in actuarial and data analytical capacity |
| Stage 3<br>Performance | Outcomes monitoring feeds payment adjustments; autonomous purchasing body established; fraud detection operational | Benefit package still partially political; not yet fully evidence-based; interoperability gaps persist | Indonesia JKN (mid-phase); Turkey post-2010 SSGSS reforms; Philippines PhilHealth advanced | Implement outcomes-linked payment; formalise intergovernmental coordination mechanisms |
| Stage 4<br>Full Strategic | All six functions as theoretically specified; cost-effectiveness drives benefit design; pay-for-performance embedded; population-level accountability | Theoretical ideal; no LMIC has fully achieved; Thailand closest but still evolving | Thailand NHSO (nearest); Estonia; New Zealand; mature OECD purchasers | Continuous evidence-based benefit review; advanced risk-adjustment; population health management |

*Pakistan's Sehat Sahulat Program is positioned at Stage 1. International policy prescriptions typically assume Stage 4 capacity. The SPCS model argues that prescriptions must be calibrated to actual capacity stage. SSP = Sehat Sahulat Program; NHSO = National Health Security Office (Thailand); JKN = Jaminan Kesehatan Nasional (Indonesia). Progression requirements in the final column reflect the minimum institutional investments required for transition to the next stage.*

The SPCS model has three implications, redefine success metrics for each stage of capacity building, establish a logical sequencing for capacity building investments, and ensure that International Agencies have strategic purchasing recommendations that are aligned with the actual purchasing capacity of LMICs, as opposed to simply transferring purchasing models from High-Capacity Contexts.

### Devolution as Structural Amplifier-The 18th Amendment Dimension

In addition to transferring health system control from the central government to the provinces through the 18th Amendment in Pakistan, the amendment resulted in two issues that are critical to the effectiveness of purchasing. Post-devolution landscape lacked both common data sets for cross provincial comparison (17) and a mechanism for the transfer of funds (20) to sustain the program implementation. Lagomarsino et al. stated that one of the primary difficulties in coordinating across agencies in federal systems arises from a lack of clarity regarding agency responsibilities. As such, the lack of clear authority also results in the inability of the agencies to coordinate effectively between the provinces. This is clearly evident in Pakistan where there has been a long-standing contest over authority between the federal and provincial governments. Provincial governments have viewed federally initiated health insurance programs as an encroachment upon the province’s new-found autonomy in the provision of health services. The issue is primarily political and will require negotiations to resolve. There also appears to be a considerable difference in perceived feasibility ratings by federal versus provincial experts concerning the implementation of health insurance programs, indicating a great deal of difference between the federal and provincial institutions. The differences in the ways federal and provincial actors perceive the feasibility of implementing health financing reforms have significant implications for policy process theory. These differences create a policy/implementation gap where the federal government expects a great deal from the provincial governments but does not provide them with sufficient resources to meet these expectations. Therefore, it creates a barrier to the effective implementation of policy (Table 6). Kingdon’s Multiple Streams Model conceptualizes how policy windows open as a function of the convergence of three elements: problem recognition, available solution, and political receptivity. A single convergence occurs in a unitary government at the national level. In Pakistan’s federal system with devolution of authority from the national to subnational levels, convergence must occur at two different levels; first at the national level where the concept and funding for an insurance policy are determined, and second at the subnational level (provinces) where the delivery of the services is implemented. The systematic differences found in this study and illustrated through data collected from federal and provincial actors regarding their perceptions of the feasibility of implementing reforms across multiple areas, indicate that there is no alignment between the federal problem and political streams and the provincial streams. The result is a unique failure mechanism, that is, while federal streams will align and create a policy window at which time policy is developed and passed at the federal level, the policy fails to be implemented as the provincial streams do not converge at the same time as the federal streams. As such, this work extends Kingdon’s model by establishing that devolved federal systems require a new multi-level convergence condition as a necessary condition for reform opportunities, and that this new condition represents a significant constraint on reform opportunities relative to those conditions assumed by single tier applications of the model.

**Table 6.** – Federal-Provincial Positioning Divergence Across Four Governance Domains.

| Policy Domain | Federal Perspective | Provincial Perspective | Structural Implication |
| --- | --- | --- | --- |
| Fiscal Responsibility | Federal bears premium cost; views sustainability as manageable under current trajectory | Provinces absorb operational and co-financing burdens not reflected in federal fiscal projections | Systematic under-estimation of programme cost at implementation level |
| Constitutional Authority | Health insurance framed as cross-cutting coordination function; federal mandate assumed | Post-18th Amendment, provinces assert full health authority; federal SSP seen as encroachment | Contested legal basis creates legitimacy deficit undermining programme stability |
| Implementation Complexity | Standardised national model seen as technically straightforward to roll out provincially | District-level heterogeneity, urban informality, and capacity gaps make standard models unworkable | Gap between policy adoption (21) and implementation failure (provincial) |
| Data & Accountability | Claims data reported upward satisfies federal monitoring requirements | Claims data does not translate into local quality feedback or service improvement | Monitoring infrastructure serves administrative compliance, not performance improvement |
| *Divergence patterns derived from Delphi expert consensus process (n=10, Rounds 1-4) and key informant interview thematic analysis (n=22). Mann-Whitney U statistics reported for federal versus provincial subgroup differences on quantitative Delphi items. Structural implications in the rightmost column are interpretive inferences from integrated qualitative and consensus data.* |  |  |  |

### Policy Implications for International Agencies

There are three major changes in how international agencies can support LMIC health financing development, based on the results of this study. Firstly, a staged capacity assessment prior to making prescriptions for strategic purchasing should occur. Prior to recommending any type of procurement strategy (such as provider performance contracting), advisory teams should assess where the system is located on the SPCS model and make recommendations based on Stage-specific investments. For example, there is no utility in recommending that a system that is unable to reliably collect data on utilization by facility engage in provider performance contracting.

Secondly, international agencies need to provide capacity-building investments in sequence before expecting functional reforms. There is a large body of evidence demonstrating that institutions are developed through investments in prior to programmatic expansions, not the other way around. This includes Tangcharoensatien et al.’s analysis of Thailand’s journey toward achieving universal health coverage and the ThinkWell report on Pakistan’s specific PHC purchasing challenges, among others (19) (22).

Thirdly, international health financing frameworks need to be revised to account for the impact of federalism on the ability to achieve health purchasing functions above Stage 1. Most health financing frameworks assume a unitary government context. However, in countries like Pakistan, Kenya (post 2010), Nigeria, and Nepal (post 2015) that have recently been devolved, health purchasing authority is contested terrain. International advisory frameworks need to include specific guidance on intergovernmental coordination mechanisms, not as optional governance enhancement options but as required structural elements of any purchasing function above Stage 1.

### Strengths, Limitations, and Directions for Future Research

This study demonstrates methodological strengths, including purposeful sampling of 22 key informants, achieving saturation among five stakeholder groups, and a Delphi process with full retention and over 75% consensus, validating qualitative results. The structured interview protocol based on Cashin & Gatome Munyua’s framework enabled comparison with international strategic purchasing evidence. However, limitations include being a single-country qualitative case study, making results analytically generalizable but not statistically representative of all low-income contexts (23). Data collected in 2022 may not reflect current conditions in Pakistan due to economic and political changes, though identified structural issues likely persist. Future research directions include: 1) Cross-national comparative studies using the SPCS model in varying contexts, 2) Longitudinal studies of Pakistan’s SSP to track institutional capacity over 5-10 years, and 3) Comparative research on health insurance purchasing in devolved federal systems to develop evidence on intergovernmental coordination mechanisms (23).

## Conclusion

The problem with strategic purchasing theory is a structural one. It depends upon institutional requirements that do not exist in systems such as Pakistan’s Sehat Sahulat Program. Therefore, its shortcomings are not caused by management decisions or political will, rather they are due to a lack of essential components (purchaser autonomy and a stable government) necessary for effective strategic purchasing.

*The main point of this paper is that strategic purchasing theory needs to include a precondition theory-a formulation of the minimum governance capacity requirements below which its prescriptions become structurally impossible to implement- and that the experiences of the Sehat Sahulat Programme in Pakistan represent the empirical data from which such a precondition theory may be developed.*

This research improves precondition theory via four major contributions (Table 7). First, the Strategic Purchasing Capacity Stages Model illustrates how purchaser sophistication develops from Stage 1 to Stage 4; thus, this model requires targeted strategic recommendations based on the purchaser’s current level of development (capacity). Second, the Sehat Sahulat program in Pakistan represents an example of a Stage 1 purchaser environment, whereas most other models interpret it as being at least at Stage 3, which demonstrates the potential for structural errors to result in avoidable failure. Third, the findings of hierarchical degradation provide an empirical base for the SPCS model indicating that administrative functions remain intact even when there are significant shortages of purchasing capacity, yet performance accountability functions disappear earliest since they require more extensive institutional infrastructure. Fourth, post 2010 constitutional devolution in Pakistan increases the difficulty of implementing purchasing reform as it creates a need for aligned political will among all levels of government (federal and provincial), demonstrating different institutional realities. This research provides additional implications beyond Pakistan; as it suggests similar problems may be present in South Asia, Sub-Saharan Africa, and post-conflict states, and therefore the necessity of developing fundamental purchasing capabilities prior to advancing aggressive programs.

*Strategic purchasing theory does not need to be abandoned. What strategic purchasing theory needs to do is recognize where it is likely to work and where it is likely to fail. And to cease providing the legitimacy for prescriptions that it cannot support in weak governance environments. That is the challenge that this paper presents to the global health financing community.*

**Table 7.**
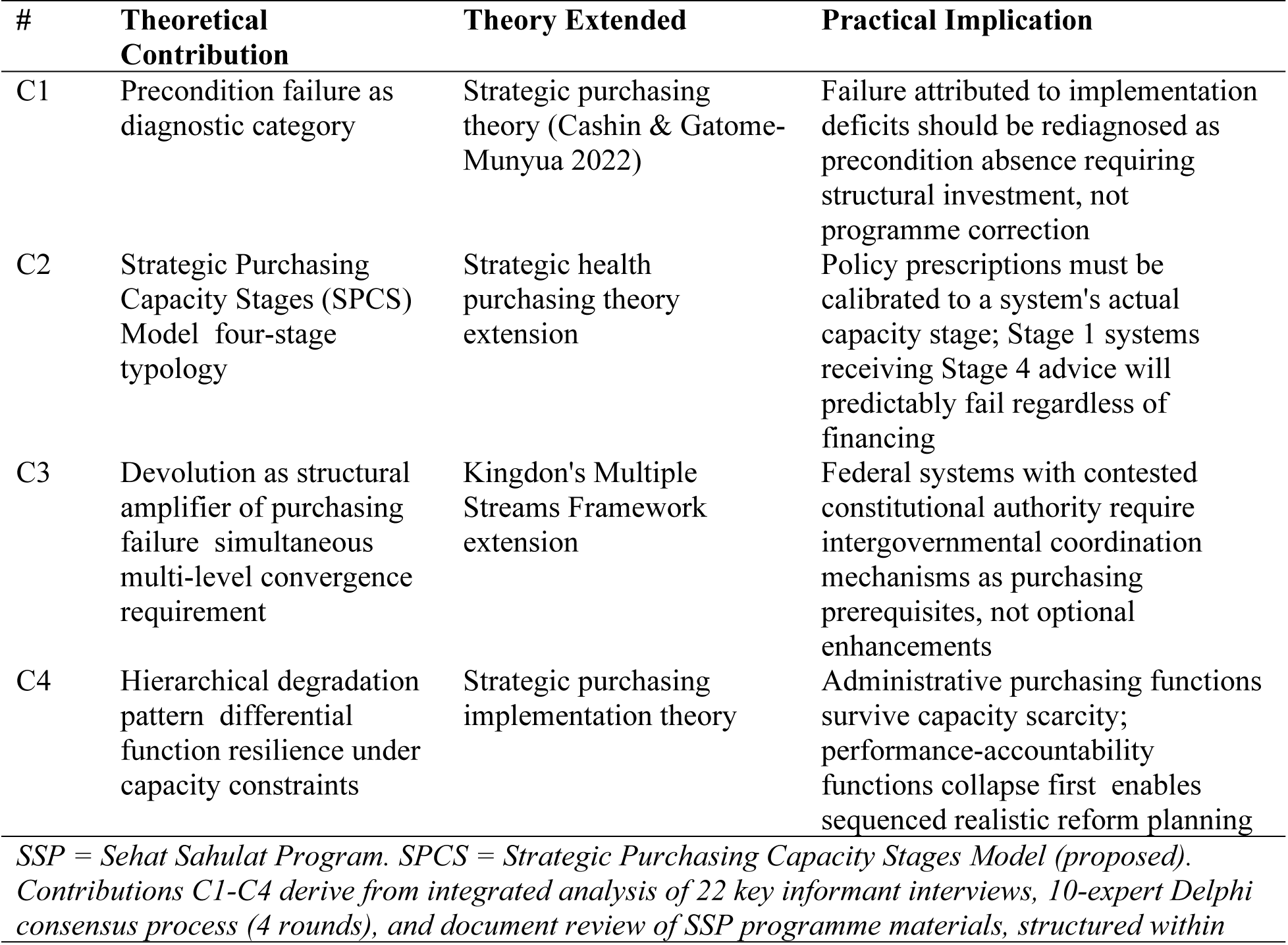
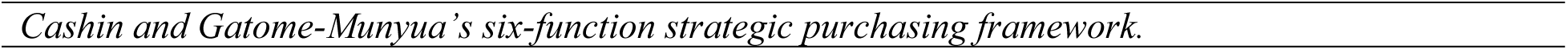
Summary of Theoretical Contributions and Practical Implications.

## Data Availability

All relevant data are within the manuscript and its Supporting Information files.

